# Ferritin and transferrin predict common carotid intima-media thickness in females: a machine-learning informed individual participant data meta-analysis

**DOI:** 10.1101/2025.04.30.25326720

**Authors:** Anand Ruban Agarvas, Richard Sparla, Janice L Atkins, Claudia Altamura, Todd Anderson, Ebru Asicioglu, Judit Bassols, Abel López-Bermejo, Hana Marie Dvořáková, José Manuel Fernández-Real, Christoph Hochmayr, Michael Knoflach, Jovana Kusic Milicevic, Silvia Lai, José María Moreno-Navarrete, Dariusz Pawlak, Krystyna Pawlak, Petr Syrovatka, Dorota Formanowicz, Pavel Kraml, Jose M Valdivielso, Luca Valenti, Martina U. Muckenthaler

## Abstract

**Background:** Iron overload promotes atherosclerosis in mice and causes vascular dysfunction in humans with Hemochromatosis. However, data are controversial on whether systemic iron availability within physiological limits affects the pathogenesis of atherosclerosis. We, therefore, performed an individual participant data (IPD) meta-analysis and studied the association between serum iron biomarkers with common carotid intima-media thickness (CC-IMT); in addition, since sex influences iron metabolism and vascular aging, we studied if there are sex-specific differences.

**Methods:** We pooled the IPD and analysed the data on adults (age≥18y) by orthogonal approaches: machine learning (ML) and a single-stage meta-analysis. For ML, we tuned a gradient-boosted tree regression model (XGBoost) and subsequently, we interpreted the features using variable importance. For the single-stage metaanalysis, we examined the association between iron biomarkers and CC-IMT using spline-based linear mixed models, accounting for sex interactions and study-specific effects. To confirm robustness, we repeated analyses on imputed data using multivariable regression adjusted for key covariates identified through machine learning. Further, subgroup analyses were performed in children and adolescents (age<18y). In addition, to evaluate causality, we used UK Biobank data to examine associations between the hemochromatosis (HFE) genotypes (C282Y/H63D) and mean CC-IMT in ∼42,500 participants with carotid ultrasound data, using sex-stratified linear regression (adjusted for age, assessment centre, and genetic principal components).

**Results:** We included IPD from 21 studies (N=10,807). The application of the ML model showed moderate predictive performance and identified iron biomarkers (transferrin, ferritin, transferrin saturation, and iron) as key features for IMT prediction. Multivariable analyses showed non-linear sex-specific relationships for ferritin and transferrin with CC-IMT: ferritin showed a significant positive association, and transferrin showed negative associations at specific ranges, both only among females. No significant associations were found between CC-IMT in those with HFE genotypes in either sex in the UK Biobank.

**Conclusion:** Our observational data show that iron biomarkers - ferritin and transferrin are non-linearly associated with CC-IMT specifically in females, while a significant causal association between the HFE genotype and CC-IMT could not be demonstrated in the UK Biobank data. We conclude that the observational associations may not only be explained by causal effects of iron on the arterial wall thickness, but also in part be driven by residual confounding factors such as inflammation.

Other: No financial support was received for this meta-analysis. The protocol for this study is registered in the PROSPERO database (CRD42020155429; https://www.crd.york.ac.uk/).

## 1. Introduction

Iron is a vital nutrient involved in numerous cellular functions; however, iron accumulation can be pathological. We have previously shown that excess iron accelerates the pathogenesis of atherosclerosis(1). Excess iron initiates redox reactions (via Fenton chemistry) that lead to oxidative damage of membrane lipids, proteins, and DNA. In addition, iron can also lead to endothelial injury, immune cell polarization, ferroptosis, and plaque destabilization, thereby, contributing to vascular dysfunction and atherosclerosis (1–3). In the blood, iron biomarkers reflective of the systemic iron status [e.g., serum iron, ferritin, transferrin, transferrin saturation (TSAT), total iron binding capacity (TIBC) and hepcidin] can be quantified. Whether these iron biomarkers within physiological limits are associated with vascular disease is of clinical relevance, but available data are conflicting.

Sonographic assessment of common carotid intima-media thickness (CC-IMT) is an indicator of arteriopathy and an independent predictor of a wide range of cardiovascular events (4, 5). Therefore, by considering CC-IMT as a surrogate marker of vascular disease we aimed to investigate its association with iron biomarkers. We also wanted to investigate the influence of sex on the association between iron biomarkers and CC-IMT. Sex-specific differences in cardiovascular disease (6) and iron metabolism (7, 8) are commonly observed. Specifically, the female sexual hormones estrogen and progesterone impair endothelial function and affect iron homeostasis by controlling hepcidin, the master regulator of iron homeostasis (9, 10). Our recent analysis in two cohorts also found a positive association between ferritin and peripheral arterial disease preferentially in females, suggesting sex-specific effects (11).

Our study also aimed to explore early vascular changes across a broader age spectrum. For example, in autopsies, atherosclerotic fibrous plaques have been detected in the aorta and coronary artery participants between 2-15 years (12). Therefore, we also included children and adolescents in subgroup analyses, to capture the onset and progression of such changes before clinical disease becomes apparent. We analysed them separately from adults since CC-IMT is strongly influenced by age and that risk factors for atherosclerosis for adults and adolescents are different.

The conclusions of previous, smaller studies that have evaluated the relationship between iron biomarkers and CC-IMT are inconsistent. Therefore, we conducted an individual participant data (IPD) meta-analysis to systematically investigate this clinically relevant question. The advantages of an IPD meta-analysis cannot be overstated: the collection and analysis of raw data from multiple studies provides a large participant-pool and thus a greater statistical power to detect smaller effect sizes (13). Working with raw data allows for standardized and flexible analyses, leading to more precise and reliable estimates.

We included a novel approach to the meta-analysis by using two orthogonal approaches that both offer unique advantages and drawbacks: machine learning (ML) and regression. Integration of ML approaches with regression-based analyses is an active area of research (14, 15), which is still being methodologically optimized. Here, we aimed to combine the strengths of predictive modelling with the interpretability and inferential capacity of traditional statistical methods. ML excels at handling complex datasets, identifying nonlinear relationships, and making predictions without requiring strong assumptions about the data. However, one of the criticisms of ML is that it lacks interpretability. On the other hand, regression analysis provides interpretability and enables testing of relationships between variables under specific assumptions. However, it may struggle with high-dimensional data and complex nonlinear patterns, as is the case with clinical data. Therefore, we used a combination of the approaches to leverage their strengths and to obtain a clear understanding of the relationships. Finally, to complement the observational analyses of the iron biomarkers, we performed a Mendelian randomization (MR) to assess the potential causal relationship between hemochromatosis (HFE) genotypes (C282Y/H63D) and CC-IMT in the UK Biobank data.

## 2. Methods

The protocol for the study is published in the PROSPERO database (CRD42020155429; https://www.crd.york.ac.uk/; Supplementary File 1). We report the study according to Preferred Reporting Items for Systematic Review and Meta-Analyses of individual participant data (PRISMA-IPD; See Supplementary File 2 for checklist (16)). We used R version 4.3.1 (17) for the data analysis and visualization (For the specific packages used, see Supplementary Table 1).

### 2.1 Literature search

We searched NLM Medline using the following string: (iron OR ferritin OR transferrin OR hepcidin) AND (atherosclerosis OR intima-media thickness). We applied filters for human studies published in English between 1^st^ Oct 1999 and 20^th^ Oct 2019 (last updated on 24^th^ Aug 2023). We used a three-step process for the study selection. In step 1, we screened the titles of the published studies and excluded records when the titles were specified as reviews or were performed on preclinical models. In step 2, we screened the abstract for human studies with one of the following keywords: iron, ferritin, transferrin, hepcidin, atherosclerosis or intima-media thickness. In step 3, we screened the full text and if studies contained data on iron biomarkers [iron, ferritin, transferrin, hepcidin, or transferrin saturation (TSAT)] and CC-IMT, they were included in the IPD. Screening of the retrieved records was done independently by two investigators (ARA, RS).

### 2.2 Data collection

We contacted the investigators (first or corresponding authors) of eligible studies with the study protocol and requested formal consent for participation (for a scheme of data request, see Supplementary Figure 1). Through subsequent contact, we asked the investigators for anonymized data on the following variables: age, gender, CC-IMT, serum iron indices (iron, ferritin, transferrin, TSAT, hepcidin), ethnic profile, and presence of comorbidities [e.g., diabetes, hypertension, chronic kidney disease (CKD), hemochromatosis, thalassemia]. For prospective studies, only the baseline data were obtained. We sent three follow-up reminders to investigators who had not responded.

From the studies for which IPD were received, we extracted the data and screened for inclusion in the meta-analysis. We piloted the data extraction by harmonizing the coding of categorical variables (e.g., gender, comorbidities) and their units of measurements, in the case of quantitative variables (Table 2). At this stage, we also compared (variables available in the received datasheets and their original publications) and identified variables common to various studies. When required, we contacted investigators again to request additional variables of interest.

Subsequently, we reextracted the data and checked the files for data integrity in three steps. In step 1, we compared the number of data participants, sex ratio, and the summary data of variables between the data file and its corresponding original paper. In the step 2, we verified the frequency distribution of continuous variables of interest in the individual data files. If required, we contacted the authors in a third step for clarifications. We excluded studies that did not clear the data integrity check.

### 2.3 Participant-level data

From the received data, all participants with an available CC-IMT measurement were selected for pooling from the selected studies. Next, we selected the commonly measured serum iron biomarkers (iron, ferritin, transferrin, and TSAT) and the demographic and laboratory variables: age, sex, body mass index (BMI), smoking status, presence of comorbidities (diabetes, hypertension, CKD, thalassemia, hemochromatosis), creatinine, hemoglobin, high-density lipoprotein cholesterol (HDLc), low-density lipoprotein cholesterol (LDLc), triacylglycerols, fasting glucose, c-reactive protein (CRP), systolic blood pressure (SBP), and diastolic blood pressure (DBP).

When discrete CC-IMT measurements were available on the right and left carotid arteries, the mean CC-IMT was calculated and used for downstream analyses (without further transformations). The data on age, sex, BMI, and the presence of diabetes, hypertension, CKD, thalassemia, and hemochromatosis were used, as indicated, in the original data files. Since the reporting units of the laboratory variables were not uniform, this required harmonization by conversion factors (e.g., mg/dL to mmol/L). Subsequently, we pooled the IPD and added our own data (11) from 323 individuals from the Heidelberg Study on Diabetes and Complications (HEIST-DiC study; https://clinicaltrials.gov, NCT03022721).

At the participant level, we applied the following exclusion criteria: 1) CC-IMT value suggestive of an atherosclerotic plaque (18) (>1.5 mm) 2) diagnosed thalassemia or hemochromatosis 3) TSAT and ferritin values suggestive of possibly hemochromatosis (19) and, 4) CRP>10 mg/dL suggesting overt inflammation (20). We also analyzed the data for missing values and imputed them for downstream analyses (described below).

### 2.4 Implementation of the ML framework

For the ML analysis, only adult participants (age≥18y) with a valid value for CC-IMT and for one of the iron biomarkers (iron, ferritin, transferrin or TSAT) were included. We converted categorical variables into numerical variables and split the data subsequently into Training and Test subsets stratified based on study (to ensure that proportion of each study in these subsets was comparable to the original dataset). In addition, to avoid information leakage between these subsets, we processed them independently with the following steps: imputation of missing values by bagged trees, removed variables with zero or near-zero variance, centering and scaling of numerical variables.

We implemented a gradient-boosted tree regression model using XGBoost (21). We used the Training subset further for hyperparameter tuning (using grid search) and performed a 10-fold cross-validation. We evaluated model performance using Root Mean Squared Error (RMSE) as the primary metric and finalized the best-performing model. We subsequently predicted CC-IMT values on the held-out Test subset and quantified the variables that are of importance for the model prediction. We also used SHAP (SHapley Additive exPlanations (22)) values to interpret the model and to assess feature contributions. For calculation of SHAP values, we used both the Training and Test subsets. The ML pipeline is shown in Figure 2b.

### 2.5 Regression analyses

For the regression analyses, since iron and atherosclerosis parameters show age-specific variations, we used data from adults (age≥18y) for the main analysis. Here, we first tested the association between CC-IMT and each of the iron biomarkers (iron, ferritin, transferrin, and TSAT) by linear mixed model regression in the complete (unimputed) data. We hypothesized that the relationship could be nonlinear, and therefore, flexibly modelled the relationship using spline regression (degree of freedom=4). In addition, since iron metabolism shows strong sex-specific differences, we specified it using an interaction term (e.g. ferritin*sex) in the models. This approach is widely accepted in epidemiological and clinical research when exploring whether the association between a predictor (in this case, ferritin, as a proxy for iron metabolism) and an outcome (CC-IMT) differs by a stratifying variable (sex). The use of interaction terms allows for formal statistical testing of effect modification by sex, rather than relying on stratified analyses alone (23). Further, to account for differences between the different datasets, we included “study” as a random effect in the model.

To further test the robustness of the observations from the previous step, we conducted multivariable regression analysis between CC-IMT and ferritin or transferrin. Here, we imputed the missing values based on the approach by Gibbs sampler (24, 25) and generated multiple imputations (n=5) from a joint multivariable linear-mixed model. For continuous variables, the method considers the relationships between all variables at once, while for categorical variables, the model assumes that they are linked to underlying continuous variables that follow a normal distribution (26). We performed the regression on the independent imputation draws, using Rubin’s rule (27) and obtained the final imputed results. We included variables from the feature importance in ML analysis as additional covariates for the multivariable regression: smoking (reference=nonsmokers), the presence of diabetes (reference=absence), age, BMI, creatinine, HDLc, LDLc, triacylglycerols, CRP, hemoglobin, SBP, and DBP. All continuous variables (except CC-IMT) were mean-centered for the analyses (28).

#### 2.5.1 Subgroup analysis

We conducted a subgroup analysis for children and adolescents (age< 18y). Here, we also used spline regression (degree of freedom=4) and used sex as an interaction with iron biomarker (as above). However, since not all studies had children and adolescents in their study population, we fitted using linear regression (without including study as a random effect).

### 2.6 UK Biobank

Further analyses were performed in the UK Biobank to determine the associations between hemochromatosis (HFE)-genotype groups and CC-IMT. UK Biobank includes ∼500,000 community volunteers aged 39-73 years at baseline assessment (2006–2010) from 22 assessment centers across England, Scotland and Wales [as described elsewhere (29, 30)]. We included participants genetically similar to the 1000 Genomes project European reference population (31) [the categorization of this population is described elsewhere (32)], with HFE p.C282Y (rs1800562) and HFE p.H63D (rs1799945) genotype data from whole exome sequencing [methods developed by Regeneron (33)]. We analyzed a subset of these participants with available carotid ultrasound data from an imaging visit starting in 2014 [n=42,299; (34)]. Four CC-IMT variables were available in the UK Biobank imaging study [at 120, 150, 210 and 240 degrees; variable IDs 22671, 22674, 22677, 22680(34)]; from these, we calculated an overall mean CC-IMT value and performed linear regression analyses to test associations with C282Y/H63D genotype groups [C282Y^(−/-)^ H63D^(+/-)^; C282Y^(-/-)^ H63D^(+/+)^; C282Y^(+/-)^ H63D^(+/-)^; C282Y^(+/-)^ H63D^(-/-)^; C282Y^(+/+)^ H63D^(-/-)^], compared to those with no mutations [C282Y^(-/-)^H63D^(-/-)^]. Models were stratified by sex, and adjusted for age, assessment centre and ten genetic principal components (to account for genetic stratification). We also performed sensitivity analyses after excluding participants diagnosed with hemochromatosis.

## 3. Results

### 3.1 Process of data collection

We identified a total of 1,032 records via literature search, of which 887 were excluded by screening the title and abstract. The full text of the remaining 145 articles was investigated, and 108 publications were selected for the meta-analysis. We contacted the authors of the selected publications, from which we received IPD data from 22 studies (IPD retrieval 20.4%)(35–55). Of these, we excluded two studies that did not clear the data integrity checks(54, 55). Additionally, we included our own data from the HEIST-DiC study (11). The flow of the literature search is shown in Figure 1 and the outline of the analysis is shown in Figure 2a.

**Figure 1.** The outline of the study and analysis

**Figure 2.** a) The flow of literature in the IPD. b) The machine learning pipeline

### 3.2 Study characteristics

Eighteen studies included in the IPD were hospital-based (11, 35, 36, 38–46, 48–52, 56) and 3 studies were population-based (37, 47, 53). Controls were part of the study population in 12 studies (35, 36, 38–40, 44–46, 49–51, 53). The characteristics of studies and variables included in the IPD-MA are shown in Table 1 and Supplementary Table 2.

**Table 1.** Characteristics of studies included in the IPD-MA Table of the characteristics of studies that provided the IPD and included in the meta-analysis.

### 3.3 Participant characteristics

The pooling of all available datasets yielded a total of 10,807 participants. From these, we excluded participants with CC-IMT value indicative of atherosclerotic plaque(18) (>1.5 mm; N=47), CRP indicative of overt inflammation (>10 mg/dL; N=167) and all conditions suggestive of an elevated iron status [possible hemochromatosis(19) (N=27), previously diagnosed thalassemia (N=131) or hemochromatosis (N=76)]. This resulted in a final pooled participant size of 10,215. Most of the participants were adults except for 4 studies(43, 45, 51, 53) which had included children and adolescents in their work (age< 18y). Eighteen studies(11, 35–38, 42–53) included male and female participants, while 3 studies(39–41) included only males in their study population. A study-wise breakdown of participant characteristics is shown in Supplementary Table 3.

#### 3.3.1 Adults

The demographics of the adult participants (N=7,523) are shown in Table 2. Here, the proportion of males (N=4,974; 66.12%) was higher than females (N=2,549; 33.88%). Overall, the males were younger, had a higher BMI and a greater proportion of smokers (p=0.009). The proportion of participants with hypertension (p<0.0001) was also higher among males; in line with this observation, the systolic blood pressure (SBP) and diastolic blood pressure (DBP) of males were higher (Table 2). On the other hand, more female participants in the cohort had CKD (44.55%; p<0.0001; Table 2). Serum iron, ferritin, and TSAT were higher, while transferrin levels were lower among males than females (Table 2; Supplementary Figures 2-5).

**Table 2.** Characteristics of adult participants. The demographics of the adult participants (age≥18y) stratified by sex are shown. For continuous variables, the summary data are shown as Median (IQR) while for categorical variables, the data are represented as N (%). P-values as calculated by Wilcoxon rank sum test (for continuous variables) or Pearson’s Chi-Squared test (for categorical variables); significant P-values are highlighted in bold. Missing data are shown as N (%). All percentages shown are based on the total number of subjects. BMI: Body Mass Index, HDLc: High-Density Lipoprotein, LDLc: Low-Density Lipoprotein, CRP: C-reactive protein, CKD: Chronic Kidney Disease, CC-IMT: Carotid Intima-Media Thickness, TSAT: Transferrin Saturation, SBP: Systolic Blood Pressure, DBP: Diastolic Blood Pressure

#### 3.3.2 Children and Adolescents

Among children and adolescents, age (p=0.131) and BMI (p=0.838) of the participants among both sexes were not different (Table 3). The proportion of smokers was higher among females than males (76% vs 24%; p=0.002). Overall, females had lower hemoglobin levels (p<0.0001). CRP levels were not different between the sexes (p=0.226). Data on the presence of comorbidities were not available for most of the participants (>98%; Table 3). With regards to the iron parameters, males had higher ferritin and transferrin levels while iron and TSAT levels were not different between the sexes (Table 3; Supplementary Figures 2-5)

**Table 3.** Characteristics of children and adolescents The demographics of the children and adolescent participants (age< 18y) stratified by sex are shown. For continuous variables, the summary data are shown as Median (IQR) while for categorical variables, the data are represented as N (%). P-values as calculated by the Wilcoxon rank sum test (for continuous variables) or Pearson’s Chi-Squared test (for categorical variables); significant P-values are highlighted in bold. Missing data are shown as N (%). BMI: Body Mass Index, HDLc: High-Density Lipoprotein, LDLc: Low-Density Lipoprotein, CRP: C-reactive protein, CKD: Chronic Kidney Disease, CC-IMT: Carotid Intima-Media Thickness, TSAT: Transferrin Saturation, SBP: Systolic Blood Pressure, DBP: Diastolic Blood Pressure

### 3.4 CC-IMT

In all studies, the CC-IMT was measured using ultrasound, but there were differences in the methods used (Supplementary Table 4). Among the adults, the CC-IMT [median (IQR); mm] of males [0.74 (0.6-0.85)] was higher compared to females [0.71 (IQR 0.6-0.82); p<0.0001; Supplementary Figure 6]. This was also the case among children and adolescents, with higher CC-IMT among males [0.41 (0.37-0.44)] compared to females [0.39 (0.35-0.43); p<0.0001; Supplementary Figure 6].

### 3.5 Missing data

We observed two types of missing values in the IPD data: 1) systematically missing variables were present across studies (since not all studies collected the variables uniformly) 2) sporadically missing values within each study. The proportion of missing values for each variable in the pooled data is shown in Supplementary Table 5. A study-wise breakdown of missing variables is shown in Supplementary Table 6 and an age-categorized breakdown is shown in Supplementary Table 7. The pattern of co-occurrence of missing values across variables is shown in Supplementary Figure 7.

### 3.6 Machine learning

The dataset used for ML contained 5,740 participants and 21 covariates. The Training subset contained 4,306 (75%) participants while the Test subset contained 1,434 (25%) participants. The baseline characteristics between the two subsets were comparable (Supplementary Table 8). The ML model demonstrated moderate predictive performance with an R2 of 0.447, indicating that it explained 44.7% of the variance in the outcome variable; the Root Mean Square Error (RMSE) and the Mean Average Error (MAE) of the model were 0.136 and 0.101, respectively. The feature importance plot shows the contribution of each variable for the prediction of the overall model (Figure 3a). On the other hand, the SHAP plot highlights how the variables and their individual values contribute to the prediction (Figure 3b). Together, these plots emphasize the importance of each variable in our ML model’s predictions. All the iron biomarkers (transferrin, ferritin, TSAT and iron) were ranked among important predictors of CC-IMT. In addition, age, sex, CRP, creatinine, LDLc, HDLc, hemoglobin, systolic and diastolic BP, triacylglycerols, and smoking were also identified as important features.

**Figure 3.** a) Variables of importance identifed in the machine learning prediciton b) SHAPley plot of the variables contributing to the model performance

### 3.7 Regression analyses

#### 3.7.1 Adults

In the analysis of unimputed data, we found that ferritin alone showed a positive effect within specific ranges [131-233 ng/mL: β=0.08, 95% CI (0.002, 0.16), p=0.046; >233 ng/mL: β=0.16, 95% CI (0.04, 0.27), p=0.008]. A significant interactive effect was also observed between females and ferritin with CC-IMT [ferritin>233 ng/mL: β=0.04, 95% CI (0.002, 0.08), p=0.037; Supplementary Table 9]. The main effects of transferrin alone were all non-significant, however negative interactions were noted for transferrin within specific ranges among females [231-263 mg/dL: β=-0.21, 95% CI (−0.43, 0.003), p=0.054; >263 mg/dL: β=-0.73, 95% CI (−1.48, 0.01), p=0.055; Supplementary Table 10]. On the other hand, none of the terms for iron or TSAT (including interactions with sex) showed statistically significant associations with CC-IMT (Supplementary Tables 11-12).

In the multivariable analyses (Tables 4-5), we confirmed potential sex-specific effects at higher levels of ferritin and transferrin. We found that ferritin alone showed a significant effect at a specific range (131-233 ng/mL: β = 0.13, 95% CI [0.02, 0.24], p = 0.038). Further, we found a positive interaction between ferritin with CC-IMT among females with CC-IMT specifically at the higher ranges (>233 ng/mL: β = 0.54, 95% CI [0.11, 0.97], p = 0.015). Although transferrin alone showed no significant effects with CC-IMT, we found significant interactions among females at specific ranges (<199 mg/dL: β= -0.49, 95% CI [-0.93, -0.07], p = 0.05; 199-231 mg/dL: β= -0.3, 95% CI [-0.56, -0.04], p = 0.052; 231-263 mg/dL: β= -0.99, 95% CI [-1.81, -0.17], p = 0.039).

**Table 4.** Multivariable regression analysis of ferritin and CC-IMT We flexibly modelled the relationship between ferritin and CC-IMT using spline regression (degree of freedom=4). In addition, we tested the associations between ferritin and CC-IMT for differences due to sex-by using an interaction term (ferritin*sex; Males as reference). All continuous variables were mean-centered for the analyses. BMI: Body Mass Index, HDLc: High-Density Lipoprotein, LDLc: Low-Density Lipoprotein, CRP: C-reactive protein, CC-IMT: Carotid Intima-Media Thickness, SBP: Systolic Blood Pressure, DBP: Diastolic Blood Pressure.

**Table 5.** Multivariable regression analysis of transferrin and CC-IMT We flexibly modelled the relationship between transferrin and CC-IMT using spline regression (degree of freedom=4). In addition, we tested the associations between transferrin and CC-IMT for differences due to sex-by using an interaction term (transferrin *sex; Males as reference). All continuous variables were mean-centered for the analyses. BMI: Body Mass Index, HDLc: High-Density Lipoprotein, LDLc: Low-Density Lipoprotein, CRP: C-reactive protein, CC-IMT: Carotid Intima-Media Thickness, SBP: Systolic Blood Pressure, DBP: Diastolic Blood Pressure.

The effect of sex alone was nonsignificant in both models. Other significant variables were age (β = 0.14, p < 0.001), diabetes (β = 0.02, p < 0.001), HDL-C (β = -0.006, p = 0.003), LDL-C (β = 0.008, p < 0.001), smoking (β = 0.016, p < 0.001 for former smokers; β = 0.019, p < 0.001 for current smokers), CRP (β = 0.01, p = 0.024), hemoglobin (β = 0.0098, p = 0.010), systolic blood pressure (β = 0.018, p < 0.001), and diastolic blood pressure (β = -0.0087, p = 0.0006); all of these significantly associated with CC-IMT (Tables 4-5).

#### 3.7.2 Children and Adolescents

In the subgroup analysis, we found no significant associations between any of the iron biomarkers and CC-IMT among children and adolescents and no notable interactions with sex (Supplementary Tables 13-16).

### 3.8 UK Biobank

Further, we analysed data from 42,299 UK Biobank participants for associations between HFE-genotypes and mean CC-IMT. Here, no statistically significant associations were detected in either males or females between CC-IMT in those with C282Y/H63D genotypes (including C282Y homozygotes) compared to those with no mutations (Supplementary Table 17). The associations remained non-significant in a sensitivity analysis, restricted to participants who were undiagnosed with hemochromatosis (N=42,193).

## 4. Discussion

To investigate the conflicting conclusions of the relationship between iron biomarkers and CC-IMT in previous studies, we conducted and report the first IPD meta-analysis. This meta-analysis includes diversely distributed studies (18 hospital-based and 3 population-based studies) and a study population comprising both adults (N=7,523) and children/adolescents (N=2,691). Although males were over-represented in the study population, the proportion of females in both age groups were sufficient to analyse sex-specific differences. The study participants showed different comorbidities e.g., diabetes, hypertension, and CKD. Therefore, this comprehensive dataset, the largest to date, allows robust exploration to test associations between iron parameters and CC-IMT across age groups and clinical settings. Our analysis approach is also novel since we used two complementary methods (ML and regression). Both, the ML and regression analyses identified iron biomarkers as important predictors of CC-IMT. Additionally, the regression analyses offered insights into sex-specific associations. Overall, the results demonstrate that elevated ferritin and reduced transferrin levels within the reference limits are significant predictors of CC-IMT, particularly in adult females.

Our findings can be interpreted from an ‘iron perspective’: ferritin stores cellular iron and is a well-established marker of body iron stores (57); transferrin carries iron in the blood and supplies all cell types with the metal. The observed association between elevated ferritin levels and CC-IMT could be interpreted as supportive of the “iron hypothesis” in atherosclerosis, which suggests that elevated iron stores contribute to oxidative stress and vascular damage (58, 59). This idea is further supported by our current finding that elevated transferrin levels (frequently observed in iron deficiency) are inversely associated with CC-IMT. A recent mendelian randomization study showing that an elevated iron status increases the risk of cardiovascular disease, specifically ischemic stroke provides additional support to this model (60). In this context, a genome-wide association study by Galesloot et al (61) showed that polymorphisms predicting higher hepcidin/ferritin ratios were associated with an increased atherosclerosis risk. Unfortunately, data on hepcidin and NTBI were not available in our study to evaluate their associations, and we suggest that future studies may consider including additional biomarkers such as NTBI and hepcidin in their analysis.

As an extension of this idea, the logical argument would be that individuals with genetic iron overload could be at a higher risk of developing cardiovascular disease (CVD). We have previously shown that patients with genetic iron overload conditions (e.g., hemochromatosis or thalassemia major) show elevated markers of vascular dysfunction [Intercellular Adhesion Molecule 1 (ICAM-1), Vascular Adhesion Molecule 1 (VCAM-1)] that correlated positively with non-transferrin-bound-iron (NTBI) in these patients (1, 2). Importantly, phlebotomy treatment of the hemochromatosis patients reverted the increased concentrations of ICAM-1 and VCAM-1 (1). To follow up on these data, we also investigated the UK Biobank for associations between mean CC-IMT and the hemochromatosis genotype (C282Y/H63D genotypes) using MR, but we did not detect significant associations. MR offers an important complement to observational analyses by reducing bias from confounding and reverse causation. The absence of an MR association contrasts with our observational findings, which suggested a potential non-linear relationship between ferritin, transferrin, and CC-IMT. These data may be explained by population studies that indicate that the HFE genotypes does not necessarily cause elevated body iron stores. For instance, the penetrance for elevated TSAT is approximately 100% for C282Y homozygotes and 37.5% for the corresponding heterozygotes (62). In addition, individuals with a homozygous C282Y HFE mutation may have received phlebotomy (62), which normalizes iron parameter and markers of vascular dysfunction. additionally, this discrepancy may indicate that the observational associations are driven, at least in part, by residual confounding, rather than a direct causal effect of iron status on arterial wall thickness.

Inflammation could be one such confounder since iron metabolism and inflammation are tightly interconnected processes. Therefore, our findings can also be interpreted from an ‘inflammation perspective’. Inflammation is known to aggravate atherosclerosis (63). Ferritin is a well-known acute phase protein, which is induced in response to inflammation. Similarly, transferrin levels are reduced in inflammatory conditions (64). Although, we have excluded individuals with CRP>10 mg/dL in our analysis, we cannot completely exclude low-grade inflammation as a driver of increased CC-IMT. Thus, the opposing associations of ferritin and transferrin with CC-IMT could also be an epiphenomenon to inflammation. However, in inflammatory states, we would expect hypoferremia due to iron redistribution into reticuloendothelial macrophages (65). But, CC-IMT was not associated with reduced serum iron levels in our analysis, providing a counterargument for an inflammatory response of ferritin and transferrin.

Biological sex is another factor that can influence iron metabolism and CC-IMT. It is well established that biological sex influences the risk for CVD (66). Although the overall prevalence of CVD is higher among males, several studies show a higher risk of mortality and morbidity in females due to CVD, particularly in the presence of common risk factors and comorbidities (67, 68). Female-specific risk factors include hormones, pregnancy and reproductive health (e.g. menstruation, pregnancy-associated disorders etc.). While the influence of female sexual hormones on iron homeostasis is known (9, 10), previous studies have not found consistent evidence linking factors such as parity, timing of menopause, duration of the reproductive period, use of hormone therapy or contraceptives with CC-IMT (69, 70). We have accounted differences due to sex by using it as an interaction term and show that ferritin and transferrin show nonlinear associations specifically among females. Our recent work also detected a positive nonlinear association between ferritin and peripheral arterial disease in certain ferritin ranges specifically in females [48–97 ng/mL: OR 14.59, 95% CI 1.6–135.93, P = 0.019; 98–169 ng/mL: OR 171.07, 95% CI 1.27–23404, P = 0.039; (11)]. Nevertheless, we were unable to adjust for effects of hormonal status or menstruation status in this study due to the unavailability of data. Further, whether these reproductive hormones also affect the production of other liver-expressed proteins, such as ferritin or transferrin is unclear.

The use of medications is another confounder affecting the interpretation of our study. Medications affect CC-IMT progression and, in some cases, iron metabolism. For example, the use of statins has been associated with a significant reduction in CC-IMT progression (15, 71). as well as lower ferritin levels (72, 73). Additionally, other commonly used drugs such as metformin, glucagon like peptide-1 receptor agonists, dipeptidylpeptidase-4 inhibitors, phosphodiesterase III inhibitors, calcium channel blockers, and antiplatelet agents also attenuate CC-IMT progression (74–76), while the effects on iron metabolism remain incompletely understood. The datasets analyzed here lacked detailed information on medication, representing a limitation of our study.

The strengths of our study include the comprehensive analysis of iron biomarkers with CC-IMT, thereby providing a rather complete picture of the role of systemic iron status. For the analysis, we applied both ML and regression in our approach. A key advantage of the ML approach is its ability to capture complex, nonlinear interactions between predictors. On the other hand, regression results are more interpretable due to their straightforward effect sizes and significance values and thus providing hypothesis-driven insights. Despite its advantages, the ML approach has limitations, including its reliance on large datasets for optimal performance and the potential for overfitting, particularly if not properly tuned. Additionally, ML models can be seen as “black boxes”, making it challenging to derive explicit causal inferences or explainability in clinical decision-making. Conversely, while regression models provide interpretable and statistically robust associations, to some extent, they assume linearity. Furthermore, regression models are susceptible to multicollinearity and require prespecified assumptions (such as defining covariates), which can limit their flexibility in identifying novel predictive patterns. In our current study, we identified variables of importance from ML and included them as covariates for the regression analyses and therefore, have integrated the two approaches. This complementary use of both methods strengthens the validity of our findings and provides a comprehensive perspective on cardiovascular risk assessment. We also recognize that the integration of ML and conventional statistics is still at an early stage; nevertheless, exploring this interface is meaningful, especially for hypothesis generation. An additional advantage of this meta-analysis is that it has achieved a good participant mix with studies from different geographical locations, a range of age groups (adults and children and adolescents), and common comorbidities such as CKD, diabetes, and hypertension. Thus, we believe that the findings are generalizable across healthy individuals, disease states and in different age groups.

The following limitations should however be considered when interpreting our findings. First, data retrieval for the IPD was incomplete, as we received only 20.4% of the published data despite our best efforts. We also had a significant proportion of missing data, both systematically across studies and sporadically within each study. Furthermore, it is essential to recognize that the data availability for these variables was not uniform across all studies; for example, we have not adjusted the models for the different treatments (e.g., medications for diabetes, lipid-lowering, dialysis for CKD, phlebotomy for hemochromatosis etc.) which could affect the CC-IMT outcome. It is worth noting that the data on additional iron biomarkers (hepcidin, NTBI) or menstrual and hormonal status were not available. In addition, the study population also does not represent all ethnicities, so we have not estimated effects due to these. While the inclusion of diverse cohorts enhances the generalizability of our findings, the heterogeneity in study designs and measurement protocols should be considered when interpreting the results. Despite the use of study-level random effects to account for between-study variability, residual confounding from unmeasured factors—such as differences in treatment regimens, comorbidities, and subtle variations in CC-IMT imaging protocols—cannot be excluded and may partly account for the observed associations. The interpretation of iron metabolism biomarkers, particularly ferritin, is complicated by significant variability in assay standardization and traceability (77), representing a limitation of our study. This variability hinders the establishment of universal reference intervals or thresholds and complicates their clinical interpretation. Therefore, the thresholds identified in our study should be considered exploratory and are not intended to serve as cut-offs for decision making. Additionally, our study is cross-sectional and does not assess the temporal relationship between the parameters; therefore, we consider that this study is for hypothesis generation and not for implying causality. Finally, since the study relied on published data, an element of publication bias cannot be excluded, as studies with non-significant results may remain unpublished and therefore, undiscovered.

## 5. Conclusion

Our observational results demonstrate that iron biomarkers (specifically ferritin and transferrin) are non-linearly associated with CC-IMT, specifically in females. However, a significant causal association between HFE genotypes and CC-IMT in the UK Biobank data were not detected. This discrepancy may indicate that the observational associations are driven, at least in part, by residual confounding factors (such as inflammation, medications), rather than a direct causal effect of iron status on arterial wall thickness. Future studies may want to consider these confounding factors alongside iron status indicators to better disentangle their specific contributions to atherosclerosis. We consider our findings exploratory to drive further research addressing the underlying mechanisms that could explain these associations.

## 6. Ethics statement

All primary studies were approved by the local Ethics committees. All participants were included in the study according to the guidelines of the local ethics committees following written informed consent to participate. The North West Multi-Centre Research Ethics Committee (Research Ethics Committee reference 11/NW/0382) approved UK Biobank, and all participants provided their written informed consent at baseline. All research was conducted in accordance with both the Declarations of Helsinki and Istanbul.

## 8. Author contributions

Anand Ruban Agarvas: Conceptualization, Methodology, Software, Formal analysis, Investigation, Data curation, Writing-Original draft, Visualization, Project administration

Richard Sparla: Investigation, Writing - Review

Janice L. Atkins (for UK Biobank): Formal analysis, Investigation, Writing-Original draft

José Manuel Fernández-Real, José María Moreno-Navarrete, Abel López-Bermejo, Judit Bassols, Krystyna Pawlak, Dariusz Pawlak, Claudia Altamura, H M Dvořáková, Ebru Asicioglu, Jovana Kusic Milicevic, Michael Knoflach, Christoph Hochmayr, Petr Syrovatka, Peter Riško, Silvia Lai, Todd Anderson: Data curation, Investigation, Writing - Review

Dorota Formanowicz, Jose M Valdivielso, Pavel Kraml, Luca Valenti: Data curation, Investigation, Writing - Review & Editing

Martina U. Muckenthaler: Supervision, Writing - Review & Editing, Project administration

## Acknowledgement

We thank the participants of all included studies. We also acknowledge the contributions of investigators and their efforts without which this data collection would not have been possible (Dr. Stefan Kopf, Dr. Rashid Merchant, Dr. Amina Abdel-Salam, Dr. Peter Risko, Dr. Tessel E. Galesloot, Dr. Dorine Swinkles, Dr. George Hahalis). We acknowledge Dr. Tiago JS Lopes’s inputs on the development of the machine learning model and Svenja Elizabeth Seide’s contributions to data preprocessing and imputation. Part of this research was conducted using the UK Biobank resource, under application 14631. We thank the UK Biobank participants and coordinators. This work used data provided by patients and collected by the NHS as part of their care and support. Copyright © (2023), NHS England. Re-used with the permission of the NHS England and UK Biobank. All rights reserved. This research also used data assets made available by National Safe Haven as part of the Data and Connectivity National Core Study, led by Health Data Research UK in partnership with the Office for National Statistics and funded by UK Research and Innovation. This study was supported by the National Institute for Health and Care Research Exeter Biomedical Research Centre. The views expressed are those of the authors and not necessarily those of the NIHR or the Department of Health and Social Care.

## 10. Data and Code Availability Statement

Part of the data (subject to data sharing restrictions) may be available from authors upon reasonable request. Codes used for data analysis and visualization are accessible here: https://github.com/griffindoc/imt

## Supplementary files

Supplementary File 1. Study protocol as published in PROSPERO database

Supplementary File 2. PRISMA-IPD checklist

**Supplementary Figure 1.** Scheme used for data requests in the study

**Supplementary Figure 2.** Rain cloud plots of serum iron categorised by age group and sex

**Supplementary Figure 3.** Rain cloud plots of serum ferritin categorised by age group and sex

**Supplementary Figure 4.** Rain cloud plots of serum transferrin categorised by age group and sex

**Supplementary Figure 5.** Rain cloud plots of serum TSAT categorised by age group and sex

**Supplementary Figure 6.** Rain cloud plots of CC-IMT categorised by age group and sex

**Supplementary Figure 7.** Figure showing the pattern of co-occurrences and instances of missing values in the dataset

**Supplementary Table 1.** List of R packages used in the analysis

**Supplementary Table 2.** Variables included in IPD-MA Table of the variables, included in the IPD with a breakdown of the number of data points, number of studies that contained a variable and the sources.

**Supplementary Table 3.** Table of the study-wise breakdown of participant characteristics

**Supplementary Table 4.** Methodological differences in CC-IMT measurement

**Supplementary Table 5.** Table of a breakdown of the proportion of missing values for each variable in the pooled data

**Supplementary Table 6.** Table of a studywise breakdown of the proportion of missing values for each variable

**Supplementary Table 7.** Table of a breakdown of the proportion of missing values for each variable categorised by age group

**Supplementary Table 8.** Comparison of baseline characteristics between the Training and Test subsets used for Machine Learning

**Supplementary Table 9.** Regression output Ferritin vs CC-IMT in adults We fitted a linear mixed model in (estimated using REML and nloptwrap optimizer) to predict CC-IMT with ferritin and sex (formula: imt ∼ 1 + ns(ferritin, df = 4) * sex). The model included study as random effect (formula: ∼1 | study). Complete (unimputed) data was used. CI = Confidence Interval

**Supplementary Table 10.** Regression output Transferrin vs CC-IMT in adults We fitted a linear mixed model (estimated using REML and nloptwrap optimizer) to predict CC-IMT with transferrin and sex (formula: imt ∼ 1 + ns(transferrin, df = 4) * sex). The model included study as random effect (formula: ∼1 | study). Complete (unimputed) data was used. CI = Confidence Interval

**Supplementary Table 11.** Regression output TSAT vs CC-IMT in adults We fitted a linear mixed model (estimated using REML and nloptwrap optimizer) to predict CC-IMT with TSAT and sex (formula: imt ∼ 1 + ns(TSAT, df = 4) * sex). The model included study as random effect (formula: ∼1 | study). Complete (unimputed) data was used. CI = Confidence Interval

**Supplementary Table 12.** Regression output Iron vs CC-IMT in adults We fitted a linear mixed model (estimated using REML and nloptwrap optimizer) to predict CC-IMT with iron and sex (formula: imt ∼ 1 + ns(iron, df = 4) * sex). The model included study as random effect (formula: ∼1 | study). Complete (unimputed) data was used. CI = Confidence Interval

**Supplementary Table 13.** Regression output Ferritin vs CC-IMT in children and adolescents We fitted a linear model (estimated using maximum likelihood) to predict CC-IMT with ferritin and sex (formula: imt ∼ 1 + ns(ferritin, df = 4) * sex). Complete (unimputed) data was used. CI = Confidence Interval

**Supplementary Table 14.** Regression output TSAT vs CC-IMT in children and adolescents We fitted a linear model (estimated using maximum likelihood) to predict CC-IMT with TSAT and sex (formula: imt ∼ 1 + ns(TSAT, df = 4) * sex). Complete (unimputed) data was used. CI = Confidence Interval

**Supplementary Table 15.** Regression output Iron vs CC-IMT in children and adolescents We fitted a linear model (estimated using maximum likelihood) to predict CC-IMT with iron and sex (formula: imt ∼ 1 + ns(iron, df = 4) * sex). Complete (unimputed) data was used. CI = Confidence Interval

**Supplementary Table 16.** Regression output Transferrin vs CC-IMT in children and adolescents We fitted a linear model (estimated using maximum likelihood) to predict CC-IMT with transferrin and sex (formula: imt ∼ 1 + ns(transferrin, df = 4) * sex). Complete (unimputed) data was used. CI = Confidence Interval

**Supplementary Table 17.** Associations between hemochromatosis HFE-genotypes and mean CC-IMT in the UK Biobank data Restricted to UK Biobank participants of European genetic ancentry. Models stratified by sex and adjusted for age, assessment centre and first ten genetic principle components. Mean of CC-IMT calculated from different measurements at 120/150/210/240 degrees.

Supplementary file 2. PRISMA-IPD checklist

