## Supplementary figures and images for "Ferritin and transferrin predict common carotid intima-media thickness in females: a machine-learning informed individual participant data meta-analysis"

### Supplementary Figure 1

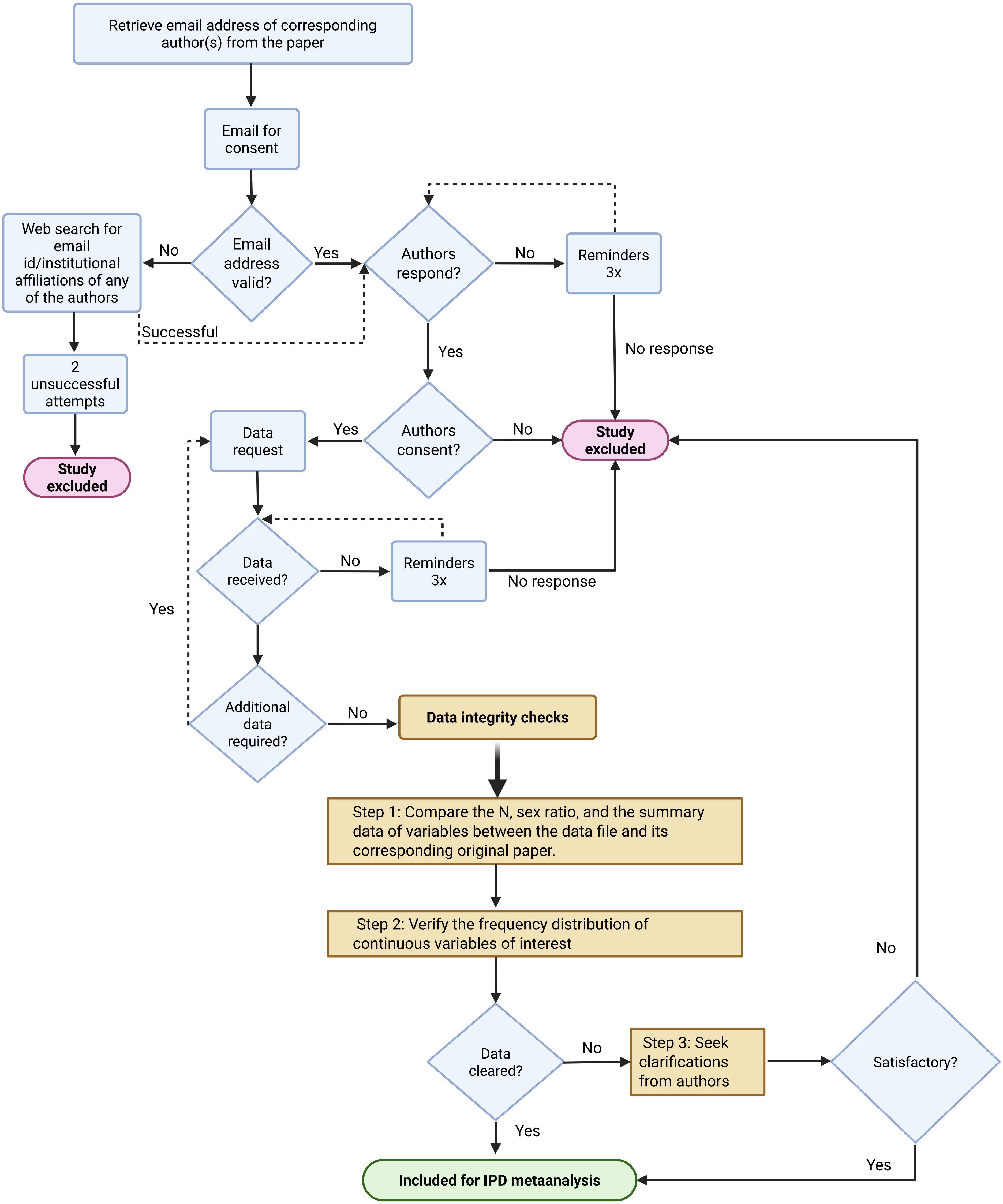
